# COVID-19 related concerns of people with long-term respiratory conditions: A qualitative study

**DOI:** 10.1101/2020.06.19.20128207

**Authors:** Keir EJ Philip, Bradley Lonergan, Andrew Cumella, Joe Farrington-Douglas, Michael Laffan, Nicholas S Hopkinson

## Abstract

**BACKGROUND:** The COVID-19 pandemic is having profound psychological impacts on populations globally, with increasing levels of stress, anxiety, and depression being reported, especially in people with pre-existing medical conditions who appear to be particularly vulnerable. There are limited data on the specific concerns people have about COVID-19 and what these are based on.

**METHODS:** The aim of this study was to identify and explore the concerns of people with long-term respiratory conditions in the UK regarding the impact of the COVID-19 pandemic and how these concerns were affecting them. We conducted a thematic analysis of free text responses to the question “What are your main concerns about getting coronavirus?”, which was included in the British Lung Foundation/Asthma UK (BLF-AUK) partnership COVID-19 survey, conducted between the 1^st^ and 8^th^ of April. This was during the 3^rd^ week of the UK’s initial social distancing measures.

**RESULTS:** 7,039 responses were analysed, with respondents from a wide range of ages, gender, and all UK nations. Respondents reported having asthma (85%), COPD (9%), bronchiectasis (4%), interstitial lung disease (2%), or ‘other’ lung diseases (e.g. lung cancer) (1%). Four main themes were identified: 1) vulnerability to COVID-19; 2) anticipated experience of contracting COVID-19; 3) wide-reaching uncertainty; and 4) inadequate national response.

**CONCLUSIONS:** The COVID-19 pandemic is having profound psychological impacts. The concerns we identified largely reflect objective, as well as subjective, aspects of the current situation. Hence, key approaches to reducing these concerns require changes to the reality of their situation, and are likely to include i) helping people optimise their health, limit risk of infection, and access necessities; ii) minimising the negative experience of disease where possible, iii) providing up-to-date, accurate and consistent information, iv) improving the government and healthcare response.

## INTRODUCTION

The COVID-19 pandemic is having profound direct and indirect impacts on health and wellbeing. In particular features of stress, anxiety and depression are being reported in populations globally(1-7), and there is evidence that a history of prior medical problems is a particular risk factor(3, 8, 9). This is of particular relevance when considering the psychological impacts of the pandemic for people with long-term respiratory conditions who are both more vulnerable to the infection(10) and more likely to have pre-existing mental health conditions(11-13).

In addition to direct health risks from the disease itself, major disruptions to normal life have occurred globally, due to approaches to reduce transmission of the virus. These have included large-scale reorganisation of healthcare systems and the introduction of measures to reduce the spread of infection, which may also be having negative psychological impacts(14). Such measures are often more burdensome for those most at risk from COVID-19 such as people with underlying health problems. In the UK people with severe respiratory conditions were among those defined as “extremely clinically vulnerable” and advised to “socially shield”, avoiding all face-to-face contact for at least 12 weeks in the first instance. All those with chronic lung conditions were defined as “clinically vulnerable” and “strongly advised” to socially distance. While the wider public, were “advised” to socially distance, including staying at home and only going outside for specific limited reasons(15-17).

Although a growing body of research has identified that mental health is being adversely impacted by the COVID-19 pandemic, limited data exist as to what exactly people are concerned about, and what their anxieties are based on. Understanding this is vital to inform our response to this burgeoning psychological crisis, as it is a necessary first step to developing information and other strategies to address and reduce distress.

The aim of this project was to systematically identify and explore the concerns of people with long-term respiratory conditions regarding the impact of the COVID-19 pandemic and how these concerns were affecting them.

## METHOD

### RESEARCH DESIGN

In this qualitative study, we used thematic analysis within a critical realist paradigm to identify and explore coronavirus related concerns of people with long-term respiratory conditions. The primary focus was to identify what the main concerns were, what was contributing to these concerns, and how these concerns were affecting people with lung disease.

We used data from an online survey conducted by the Asthma UK and British Lung Foundation (AUK-BLF) partnership, between 1st and 8th of April 2020. This was during the 3^rd^ week of national social distancing measures and advice, introduced by the UK government on 23^rd^ March 2020. This survey included the question “What are your main concerns about getting coronavirus?” and the free text responses to this constitute the data analysed here.

Quantitative data from this survey, relating to the impact of the measures to reduce risk of COVID-19, will be reported elsewhere. These were reported separately given the breadth of issues covered in the whole survey, and to allow for adequate space for the qualitative analysis of the topic presented here.

Although there are multiple coronaviruses, given the widespread impact and disruption caused by SARS-CoV-19, the virus that causes the disease COVID-19, the term ‘coronavirus’ is often used interchangeably with COVID, and COVID-19, by members of the public to refer to the disease, the virus, and the pandemic.

### RECRUITMENT & DATA COLLECTION

The survey was distributed via the AUK-BLF Partnership’s mailing lists and websites, and advertised through social media including Facebook, Twitter, Instagram and LinkedIn. 9,516 responses were collected; after excluding blank responses to the study question, 7,039 responses were analysed. Responses were anonymous; however, basic demographic and respiratory condition specific data were collected to contextualise responses (see table 1).

**Table 1:**
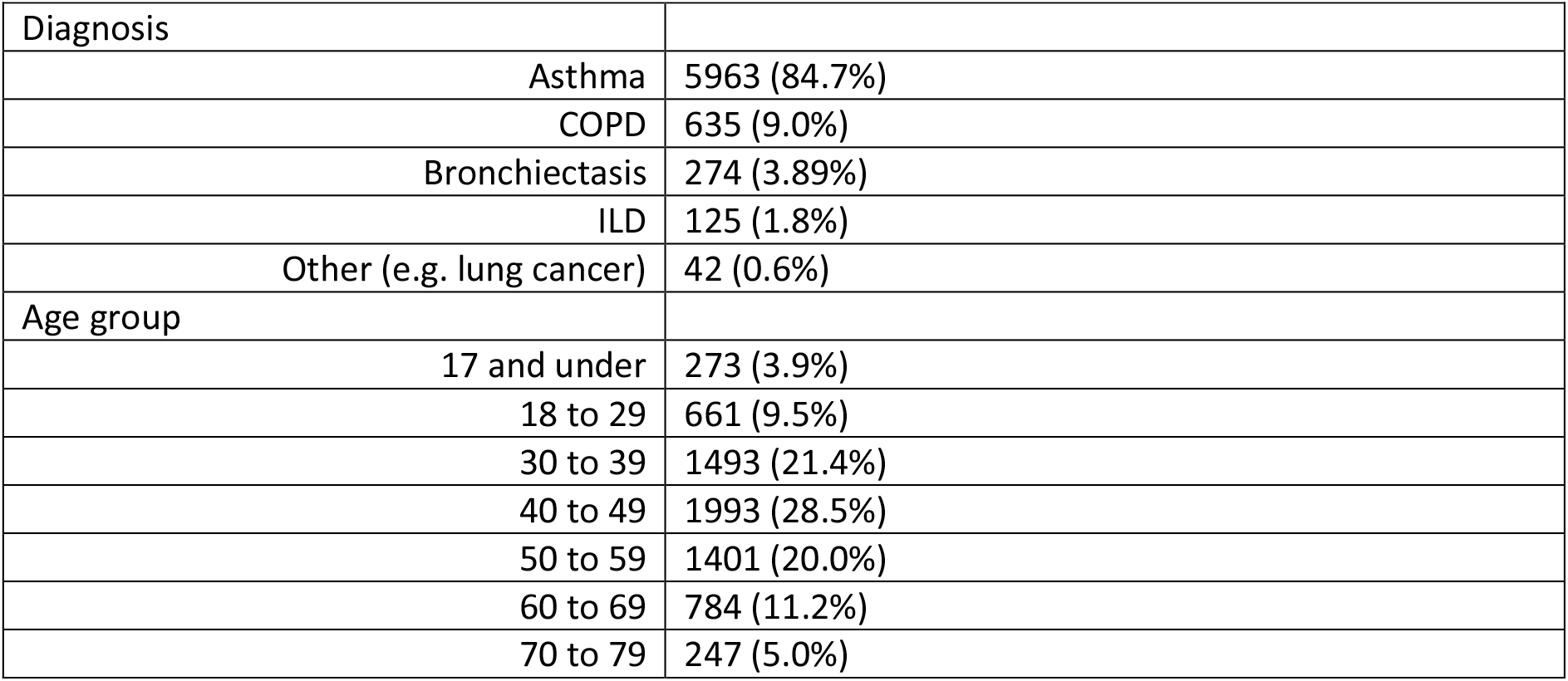
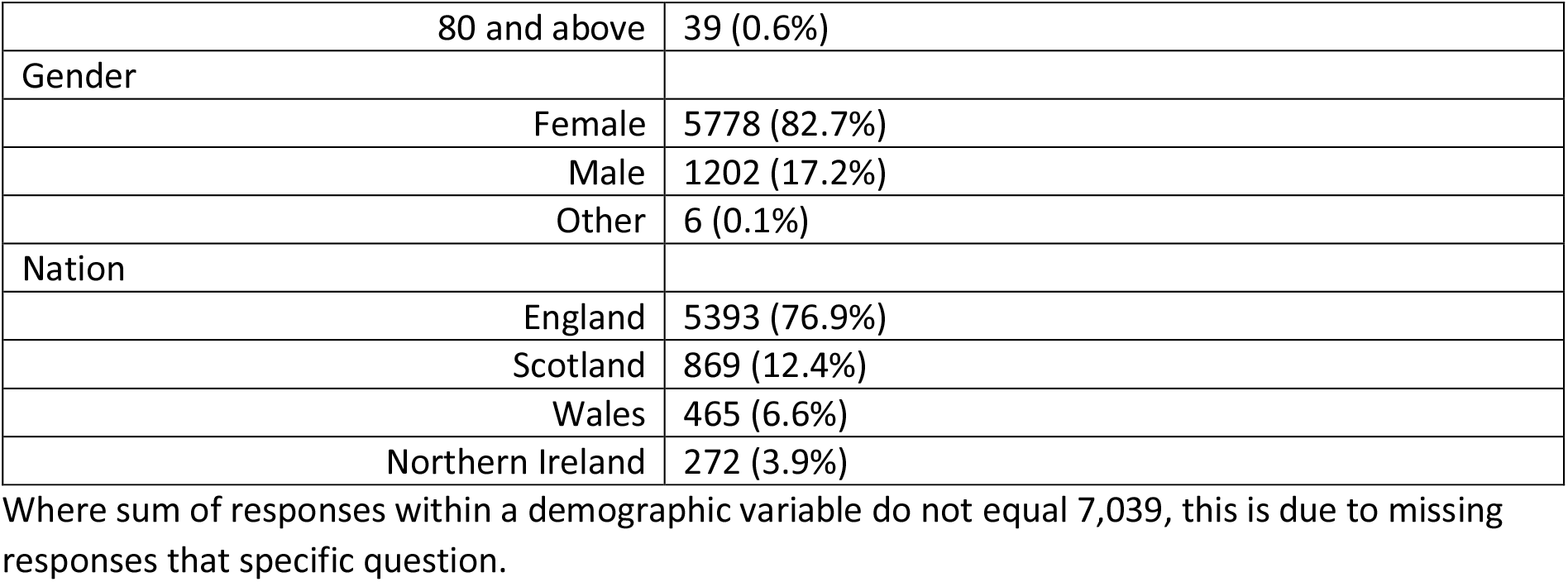
Respondent Characteristics.

| Diagnosis |  |
| --- | --- |
| Asthma | 5963 (84.7%) |
| COPD | 635 (9.0%) |
| Bronchiectasis | 274 (3.89%) |
| ILD | 125 (1.8%) |
| Other (e.g. lung cancer) | 42 (0.6%) |
| Age group |  |
| 17 and under | 273 (3.9%) |
| 18 to 29 | 661 (9.5%) |
| 30 to 39 | 1493 (21.4%) |
| 40 to 49 | 1993 (28.5%) |
| 50 to 59 | 1401 (20.0%) |
| 60 to 69 | 784 (11.2%) |
| 70 to 79 | 247 (5.0%) |

|  |  |  |
| --- | --- | --- |
|  | 80 and above | 39 (0.6%) |
| Gender |  |  |
|  | Female | 5778 (82.7%) |
|  | Male | 1202 (17.2%) |
|  | Other | 6 (0.1%) |
| Nation |  |  |
|  | England | 5393 (76.9%) |
|  | Scotland | 869 (12.4%) |
|  | Wales | 465 (6.6%) |
|  | Northern Ireland | 272 (3.9%) |

Where sum of responses within a demographic variable do not equal 7,039, this is due to missing responses that specific question.

The surveys had been carried out by AUK-BLF with the primary purpose of informing and developing the charity’s policy to assist its beneficiaries and no individual participant-identifying data were shared with academic partners. All participants in surveys consented to the use of their responses for analysis and publication and following the charity’s information governance process additional external ethical approval was not deemed necessary.

### DATA ANALYSIS

Responses and relevant demographic and disease specific information were used. KP and BL conducted a thematic analysis as described by Braun and Clarke(18). This included initial open inclusive coding which were then refined, using the context from initial readings. Initial clustering of codes into meaningful units, then preliminary themes, was done independently by KP and BL, who then came together to discuss, refine, reorganise and name the agreed themes. Themes were reviewed with the co-authors. Participant validation of the themes was not possible given the nature of the research methods employed. Quotations reported below are followed in brackets by the respondents’ gender, primary respiratory condition, nation of residence, and age group in years.

## RESULTS

The responses suggest that the coronavirus pandemic has created wide ranging and profound concerns for people with long-term respiratory conditions in the UK, far outreaching becoming personally infected. These concerns were already negatively impacting mental health, through increased stress and anxiety. Vivid descriptions illustrated the severity of these impacts:

> *‘I’m petrified to be honest. Terrified even’* (Female, COPD, Wales, 50-59)
>
> *‘my anxiety has risen feeling like my throat is closing and palpations, along with COPD feeling panicky’* (Female, COPD, Scotland, 40-49)
>
> *‘Nobody seems to care and I’m tearful and losing sleep’* (Female, asthma, England, 40-49)

Four key themes were identified, which were concerns about (1) Vulnerability to COVID-19 (most dominant theme); (2) anticipated experience of contracting COVID-19; (3) Wide-reaching uncertainty; and (4) Inadequate national response. There was also substantial interaction between these themes, with concerns around ‘vulnerability to COVID-19’ (theme 1), being closely linked to, yet distinct from, concerns regarding the ‘anticipated experience of contracting COVID-19 (theme 2)’, partially driven by wide-reaching uncertainty (theme 3) and the perceived inadequacy of the national response (theme 4, see Figure 1).

**Figure 1:** Thematic Map.

### THEME 1: VULNERABILITY TO COVI-19

> *‘I know it would kill me’* (Female, COPD, England, 70-79)

Immediate personal vulnerability to severe disease and death was very commonly described, *‘I know it would kill me’* (Female, COPD, English, 70-79). Concerns were also expressed regarding vulnerability to longer-term impacts on both physical health, *‘Permanent further lung damage’* (Female, COPD, England, 60-69), and mental health, *‘PTSD if I recover from being on a ventilator’* (Female, COPD, England, 40-49), which may cause *‘even poorer quality of life’* (Female, bronchiectasis, England, 30- 39).

#### Pre-existing health conditions and Denial of Care

Vulnerability amongst respondents was primarily attributed to their pre-existing respiratory condition *‘That I will die from it due to my asthma’* (Female, asthma, England, 30-39), and *‘That it will be fatal. I have lung fibrosis. I’m not panicking, I’m being realistic’* (Female, pulmonary fibrosis, Scotland, 60-69). However, non-respiratory co-morbidities were also commonly mentioned, *‘I have asthma diabetes and I’m obese, I feel like I’m a sitting duck’* (Female, asthma, England, 50-59).

This was compounded by frequently reported concerns regarding being denied care, due to having a pre-existing medical condition, *‘health officials will let me die because I have asthma I won’t be seen as of any value’* (Female, asthma, England, 30-39), or due to age, *‘because of my age, I will be written off’* (Female, COPD, England, 70-79).

Particularly notable was the emotive nature of the language used to describe these concerns:

> *‘They will consider I’m not worth trying to save. So I won’t see my family ever again’* (Female, ILD, England, 70-79)
>
> *‘I feel forgotten’* (Female, bronchiectasis, England, 30-39)

Concerns regarding denial of care were presented as resulting from discrimination (based on age or pre-existing medical conditions, as above); inadequate resources leading to rationing (see Theme 4 below); or a combination of these factors.

A small number suggested the potential denial of care was concerning but possibly justified, *‘its totally understandable and ethical and I’d gladly give my life to someone more likely to survive’* (Female, COPD, Wales, 60-69). However, the majority did not agree with this and were concerned about *‘not being treated fairly’* (Female, bronchiectasis, England, 40-49):

> *‘I have a manageable condition that is under control with medication, every life should count and every life should ideally be given the same chance of respiratory treatments without discrimination’* (Female, asthma, England, 50-59)

#### Exposures

Respondents had concerns about ongoing exposures, particularly for those whose profession or personal circumstances made further reductions in exposure impossible, *‘My husband bringing it into the house*…*we are sleeping in separate rooms’* (Female, asthma, England, 60-69). Coronavirus was described as *‘the invisible enemy’* (Female, asthma, England, 50-59) and respondents were worried about unwittingly passing the virus on before they developed symptoms:

> *‘Patients not tested until showing symptoms and more evidence coming through that half of transmissions could be from a symptomatic people. I’m absolutely petrified of this invisible disease’* (Female, asthma, England, 40-49)’

Responses discussing ‘Key Workers’ frequently cited concerns about working conditions that put them at increased risk, including inadequate PPE, which in turn linked with their perceived vulnerability (theme 1):

> *‘As an NHS worker, our work is extremely unsupported, we feel like being treated like super humans who need no protection, it is scary’* (Female, asthma, England, 30-39)
>
> *‘I am a front line worker we have no PPE and no test for corona virus. Are the governments frightened to test us in case they have to send us all home to self-isolate’* (Female, asthma, Scotland, 60-69)

#### Concerns for loved ones

Vulnerability related to impaired capacity for caring roles was common, including caring for children

> *‘Who will look after My son who has learning disabilities if I don’t survive’* (Female, COPD, Wales, 50- 59), vulnerable or elderly relatives, *‘Who would care for my husband, who has Parkinson’s disease, and can no longer care for himself’* (Female, asthma, Wales, 60-69), and pets *‘What will happen to my dog. He is blind, deaf & diabetic’* (Female, other, England, 70-79).

Many were concerned about passing an infection to family members, *‘Losing my life or one of my family members catching it from me and losing their life’* (Female, asthma, England, 18-29), and beyond the family unit, *‘That I get the virus and take it to work and it kills vulnerable people’* (Female, asthma, England, 30-39), leading to death.

#### Accessing necessities

Access to food and medication and the risk of virus exposure when buying, collecting or receiving them were frequent topics of concern for respondents. Many supermarkets had implemented priority online shopping for vulnerable people, but access to delivery slots remained a challenge for some respondents, particularly if they hadn’t received a shielding letter:

> *‘I’ve shopped online with Tesco for years, but now, because I’ve not had a letter from the NHS, I can’t get a slot… I might just starve to death!!’* (Female, bronchiectasis, England, 70- 79)

Others found that prescriptions were being restricted or *‘taking a lot longer than usual’* (Female, asthma, England, 20-29) to receive.

#### Work and finances

Financial issues from being unable to work were often mentioned, despite the government’s furlough scheme. This was often linked to concerns for looking after one’s family:

> *‘I’m scared if I work and I get it my lungs won’t be able to survive it and I leave 4 children without a mother. If I don’t work I’m struggling to feed them and keep a roof over our heads we can’t survive without an income’* (Female, asthma, England, 30-39)

The uncertainty of future and ongoing employment were other key issues. Those continuing to work also had reservations, with perceptions that employers were not being considerate:

> *‘The lack of understanding from my line manager about asthma and the impact catching Coronavirus could have, I have asked about ensuring social distancing at work but not been accommodated. I work in a NHS microbiology laboratory too’* (Female, asthma, England, 40- 49)

### THEME 2: ANTICIPATED EXPERIENCE OF CONTRACTING COVID-19

> *‘It sounds an awful way to die’* (Female, England, COPD, 70-79)

The thought of experiencing the disease was a major concern, including symptoms, psychological distress, and isolation due to infection control measures.

#### Symptoms

Experiencing the symptoms of the disease was frequently mentioned, and in particular, *‘not being able to breathe’* (Female, asthma, England, 50-59), *‘struggling to breathe’* (Male, asthma, England, 30- 39) and *‘fighting breathlessness’* (Female, asthma, England, 60-69). Concerns regarding COVID-19 symptoms were often explained, and possibly exacerbated, by previous personal comparable experiences:

> *‘I’ve been hospitalised several times with pneumonia and asthma. The distress I felt at those times keeps replaying in my mind because of hearing about people on ventilators. I’m terrified of not being able to breathe and dying. I feel the breathing difficulties from coronavirus would be even worse than when I’ve been admitted to hospital in the past’* (Female, asthma, Wales, 40-49)

#### Being alone in Hospital

One of the most concerning aspects of the disease centred on separation from loved ones and having to experience the disease alone. This included being alone in hospital, *‘Having to go to hospital without a parent’* (Female, Asthma, English, ≤17). The possibility of respondents, or their relatives, dying alone was a common concern; this included worrying about not being found after death for some respondents:

> *‘I’m not scared of dying, but I don’t want to die alone, as is being reported’* (Female, bronchiectasis, English, 70-79)
>
> *‘It’s very possible that I could end up dying alone, unnoticed, in my flat, because I’m an international student’* (Female, asthma, England, 30-39)
>
> *‘That I will die and I won’t be found for weeks or months as my family and friends can’t visit me’* (Male, asthma, England, 50-59)

#### Death

Concerns regarding death included the circumstances surrounding death, beyond being alone. For example, *‘Dying a painful death’* (Female, COPD, England, 60-69), *‘dying gasping for air’* (Female, asthma, England, 60-69) and *‘sounds an awful way to die’* (Female, COPD, England, 70-79). The place of death was also important to respondents; they wanted to avoid dying *‘somewhere like Excel’* (Female, ILD, England, 70-79), in a *‘4,000 bed hospital’* (Female, asthma, England, 40-49) or an *‘empty cubicle’* (Female, lung fibrosis, Wales, 60-69).

After death, there were concerns about not having a funeral, *‘they won’t be allowed to see me in hospital or have a funeral’* (Female, asthma, England, 50-59) or being unable to attend the funeral of a loved one. Media depictions were raised, and frustration shown at the negativity of media coverage:

> *‘The news and media makes you feel as if you will definitely die if you have Coronavirus and underlying health issues like asthma. This does not help with mental health and anxiety’* (Male, asthma, England, 30-39)

#### Treatment

Being admitted to hospital was an independent concern for respondents, regardless of symptoms or severity of illness experienced. Some of the reasons given for this were *‘being helpless…with no privacy or independence’* (Female, asthma, England, 50-59) and receiving treatments (e.g. sedation, intubation, intensive care admission) associated with critical illness, *‘Becoming very ill and having to be intubated’* (Female, asthma, England, 40-49), *‘having to go into intensive care again’* (Female, COPD, England, 60-69). Some were concerned that there is *‘no medicine for it’* (Female, asthma, Northern Ireland, <18).

### THEME 3: WIDE-REACHING UNCERTAINTY

> *‘When will it end’* (Female, asthma, England, 50-59)

Participants reported uncertainty in relation to multiple key aspects of their lives, including to the disease, *‘That it is an unknown illness’* (Female, bronchiectasis, England, 40-49) their long-standing health conditions, the current situation and the future.

A key challenge was differentiating the symptoms of their pre-existing respiratory condition from symptoms of COVID-19, particularly given the concurrent allergy season:

> *‘I worry that I will catch it and struggle to breathe and won’t know if the cause is an anxiety attack, asthma attack, coronavirus or a mix of all 3’* (Female, asthma, Scotland, 30-39)

Uncertainty was also expressed regarding management of underlying health conditions, including how to manage inhalers and steroids, *‘conflicting advice on management I*.*e. to use steroids or not’* (Male, asthma, England, <18). A perceived lack of control and *‘a feeling of chaos’* (Female, bronchiectasis, England, 60-69) was central to expressed uncertainty; without control, certainty was unachievable:

> *‘I need to feel in control of my own destiny, but like the rest of us, I have no say in the matter’* (Male, COPD, England, 50-59)

Many were concerned about not having a voice, and therefore no control or certainty:

> *‘I would have liked to have been given the opportunity to discuss relevant treatment if I was to contract coronavirus. DNR is my wish if quality of life would be more effected than my current position’* (Male, COPD, England, 50-59)

The uncertainty of the future for individuals and for society was mentioned, *‘When will it end’* (Female, asthma, England, 50-59). This included what happens when the response changes, *‘I’m worried that when lock down ends the virus will come back with a vengeance’* (Female, bronchiectasis, Wales, 30- 39). Some were keen to know when a vaccine would be ready, as it was seen to be the most viable exit strategy:

> *‘For me, life can’t go back to normal until there is a vaccine but when will that be?’* (Female, asthma, Scotland, 40-49)

These uncertainties were frequently described as being drivers of psychological distress (Theme 3), and often driven by the perceived inadequacy of the government’s response and health service provision preparedness.

### THEME 4: INADEQUATE NATIONAL RESPONSE

> *‘Government are crap’* (Male, asthma, England, 50-59)

The inadequacy of the government, health service and public response was a common cause for concern. Some people felt forgotten about; that nobody cares:

> *‘No shielding letter and have to work in a patient facing role. Nobody seems to care and I’m tearful and losing sleep’* (Female, asthma, England, 40-49)

#### Government

The government response was frequently described as slow and confused, *‘Totally conflicting messages on this subject’* (Female, COPD, England, 70-79) with some being more direct, *‘Government are crap’* (Male, asthma, England, 50-59). Social distancing measures were frequently mentioned as being vital yet delayed and inadequately implemented, *‘Government did not act quickly enough nor are they strict enough with the rules’* (Female, asthma, Scotland, 50-59). Conflicting messaging caused confusion, and increased uncertainty (Theme 3).

Having not received a letter from the government advising ‘social shielding’ was a concern for many, due to the resultant risk exposure. This was particularly confusing for those who met a different set of criteria for shielding given by AUK-BLF:

> *‘The mixed information about whether I should be shielding. I haven’t had a letter but I fit criteria on BLF and some on Asthma UK’* (Female, asthma, England, 40-49)
>
> *‘I’m terrified that my asthma isn’t seen as severe enough as I haven’t had a letter or any contact to say I should shield or stay at home. I am a primary school teacher so have to attend work and I’m at a loss of what to do. I don’t feel I’ve had any support or guidance to make any decision about shielding’* (Female, asthma, England, 20-29)

A lack of testing was also raised repeatedly, *‘I am concerned that more people have not been tested’* (Female, COPD, England, 70-79).

#### Health Service

The level of preparation of the health service was often mentioned. Lack of health service capacity was a frequent concern, which in turn contributed to concerns regarding denial of care (see Theme 1), and propagated uncertainty (Theme 3). Potential gaps in provision were noted regarding multiple key aspects of care:

> *‘I am afraid I won’t be able to go to hospital because there won’t be any beds left, or I won’t be able to get the treatment I need because of lack of ventilators and doctors’* (Female, asthma, England, 40-49)

Related issues with accessing healthcare included not being treated quickly enough due to long waiting times to get through to the GP or 111 and for ambulances to arrive. This concern was heightened for those living in rural areas, with access to healthcare and social support both perceived as being more difficult:

> *‘I live in a very remote area and I don’t believe the necessary level of support will be available if I need it’* (Male, asthma, Scotland, 50-59)

The perceived inability of the health service to respond to COVID-19 was often based on the opinion that the NHS was struggling before the current crisis. The government were often presented as being wholly to blame for failings, including inadequacies in the health service; NHS staff were criticised or blamed far less often:

> *‘The fact over ten years of austerity has left our NHS unable to cope on a day to day basis before Coronavirus took hold’* (Male, asthma, England, 30-39)
>
> *‘I don’t trust that the government has put the right measures in place to protect NHS frontline staff and I don’t trust I would receive help in time if I got ill’* (Female, asthma, England, 40-49)

These opinions were frequently supported with personal experience, *‘my past experiences with our 1 hospital are not great I worry about the care they could give’* (Female, bronchiectasis, England, 40-49), and extended beyond the immediate threat, including provision for non-COVID-19 issues:

> *‘cancer recurrence…By time I get app concern is may have widespread mets and nothing can be done or such long waiting lists due to back up of patients I will not get treatment’* (Female, asthma, England, 50-59)

Of note, despite the overwhelmingly positive way in which the Nightingale hospitals were presented in the media, not all respondents were convinced:

> *‘I’m also concerned I may be treated in a temporary hospital, by inexperienced staff, using second-rate ventilators’* (Female, asthma, England, 40-49)

Though the response of the government and health service were most commonly cited sources of concern, the behaviour and actions of other members of the public was also raised, *‘All the idiots out there not following govt guidelines’* (Male, ILD, England, 50-59).

## DISCUSSION

This study identified that people with long term respiratory conditions have wide ranging and profound concerns regarding the impact of COVID-19. Anxieties regarding their own and others’ vulnerability to COVID-19 and about what the experience of contracting the disease would be like were exacerbated by perceptions of an inadequate national response to the pandemic, and wide-reaching uncertainty regarding how to deal with their situation now and in the future.

### Significance of the findings

Previous studies have identified high levels of stress and anxiety related to the COVID-19 pandemic across the world (1, 2, 8, 9, 19), particularly in people with existing medical conditions (3, 8, 9). Research exploring the specifics of these concerns has so far been limited, although what does exist has identified similar topics to the themes identified in our present work. Regarding vulnerability (theme 1), one study identified that factors associated with ‘extent of potential harm’, for example advanced age, were related to more detrimental impacts on psychological well-being (4). Similarly, concerns regarding becoming infected (self or family members) have been associated with detrimental mental health impacts (1, 7). Regarding ‘wide reaching uncertainty’ (theme 3), one study identified that access to specific up-to-date accurate health information has been associated with lower levels of COVID-19 related stress and anxiety(1), which may explain the concerns found in this study regarding theme 3, and ‘conflicting and inaccurate’ health related information. Our findings are also consistent with outbreaks of other diseases, in which social distancing or quarantine type measures were implemented. In a rapid review of the psychological impacts of quarantine, Brooks et al. (2020) identifies ‘stressors’ that were also identified in our study. These include duration of quarantine, fears of infection, a sense of isolation, inadequate supplies, and inadequate information(14).

The findings of this study are important as the COVID-19 pandemic is likely to continue to pose a threat to people with long-term respiratory conditions. The concerns raised by respondents were, in the vast majority, well-reasoned and based in their lived experience. As such, addressing these concerns requires concrete changes to reality, rather than just the participants’ perceptions of these stressors. Importantly, certain aspects of these concerns are potentially modifiable, such as the adequacy of the government and health service response, while other factors such as vulnerability related to age are not so. Although the long-term respiratory conditions reported by participants in the survey are not curable, optimisation of care and self-management would be likely to reduce individual vulnerability (20). This poses challenges, especially in the context of disruptions to normal health service provision, and issues relating to people who are clinically vulnerable to COVID-19 accessing medication and food. This is particularly the case in respiratory disease where even prior to COVID-19 there were significant areas of inadequate provision of care and unmet need (21-25).

Uncertainty regarding personal vulnerability due to pre-existing long-term respiratory conditions may have been increased due to a lack of clarity regarding who was defined as being at clinically vulnerable, or extremely clinically vulnerable, due to existing health conditions. This inevitably posed a challenge for government messaging, given the initially limited information on relative risk for different groups. Moving forward, with better epidemiological data, clearer messaging should be possible.

An important finding is that concern about the experience of developing COVID-19, appears in part to be driven by having previously had comparable symptoms. Thus, anxiety about COVID-19 for people with respiratory conditions reflects their experiences of exacerbations of existing lung disease. It follows from this that efforts to optimise delivery of evidence-based self-management(26-28) for underlying lung conditions may be useful and supports recommendations that these should be prioritised during this lockdown period(29, 30).

Anxieties about the preparedness and ability of healthcare and other systems to cope is understandable given widespread reporting of objective failings such as delayed introduction of lockdown measures, test trace and isolate systems, and the provision of personal protective equipment(31, 32). However, it was clear from some responses that this had translated into unjustified anxiety that individuals might, for example, be denied care as a matter of policy. Concerns about health service preparedness may have contributed to a reluctance to seek healthcare for other conditions leading to delayed presentations, e.g. for acute exacerbations of asthma or COPD.

### Methodological issues

This study has a number of strengths. To our knowledge it is the first large qualitative study to identify and explore the concerns of people with long term respiratory conditions regarding COVID-19. By identifying specific areas and topics of concern, this study provides valuable information to guide and shape interventions and policy decisions. In addition, the large sample size, with representation of individuals across age groups, genders, UK nations, and multiple long-term respiratory conditions ensures that both breadth and depth of analysis were possible.

Some limitations should be noted. Although the survey included a large number of respondents with diverse demographic characteristics, it is unlikely that the sample is representative of all people in the UK with long-term respiratory conditions. Of note, the high proportion of women, and people with asthma (Table 1), suggests these groups are overrepresented. Due to the use of AUK-BLF mailing lists and social media for dissemination the survey link, respondents may also be more engaged with their condition than the average person. Additionally, though our study does not compare concerns between genders, female gender has frequently been reported as being related to increased levels of reported anxiety and depression in relation to COVID-19(1, 2, 8, 9), though not in all studies (4). This is a point of note, given due to the higher representation of females in our sample. Furthermore, a degree of selection bias is likely to exist, as surveys were completed online, necessitating a degree of computer literacy. People without concerns are less likely to be represented in these data, as they were drawn from responses to the question *“What are your main concerns about getting coronavirus?”*, rather than a more neutral wording. This was because the purpose of the survey was to help AUK-BLF identify concerns among their beneficiaries to support internal decision making. Finally, more information on participants would have been useful, for example socio-economic status, previous health status and comorbidities, ethnicity and levels of anxiety.

## CONCLUSIONS

Both COVID-19 and the measures taken to contain it disproportionality effect people with long term respiratory conditions. Identifying and exploring their concerns is vital in the development of effective strategies to mitigate the psychosocial impacts of the pandemic. Key approaches are likely to include i) Helping people optimise their health, limit risk of infection, and access necessities; ii) minimising the negative experience of disease where possible, iii) Providing up-to-date, accurate and consistent information, iv) Improving the communication and delivery of national response.

## Data Availability

The datasets generated and analysed during the current study are not publicly available as they contain identifiable and personal information, for which consent for sharing was not taken.

## DECLARATIONS

### Ethics approval and consent to participate

The surveys had been carried out by AUK-BLF with the primary purpose of informing and developing the charity’s policy to assist its beneficiaries and no individual participant-identifying data were shared with academic partners. All participants in surveys consented to the use of their responses for analysis and publication and following the charity’s information governance process additional external ethical approval was not deemed necessary. Confirmation from the AUK-BLF Director of External Affairs included in submission.

### Authors’ contributions

All authors contributed to the conception of this study. AC, JFD, ML led the data collection. KP and BL conducted the analysis and wrote the first draft of the manuscript. All authors reviewed, revised and contributed to further drafts, and approved the final manuscript for submission.

## Acknowledgements

The authors would like to thank all people who participated in the survey, and the team at the BLF and AUK.

## Authors’ information (optional)

KP is a respiratory registrar (doctor in specialist training) (8 years post qualification), currently completing his PhD at Imperial College. He has training in qualitative research methods including thematic analysis.

AL is a medical doctor completing Internal Medical Training (3 years post qualification). He has completed training in qualitative research methods.

